# The influence of sleep on emotional and social recognition memory and gist abstraction in children

**DOI:** 10.64898/2026.08.18.26360653

**Authors:** P.T. Meyer-Jajkov, E.-M. Kurz, F.M. Höpfner, Z. Tunçel, L. Hebborn, J. Paetow, K. Kölle, H.-V.V. Ngo-Dehning, A. Conzelmann, A. Prehn-Kristensen

**Author notes:** PTMJ and EMK share first authorship. HVVND, APK and AC share last authorship. **Corresponding authors** Alexander Prehn-Kristensen, Annette Conzelmann.

## Abstract

Sleep is proposed to have a beneficial effect on the consolidation of memories and gist abstraction in adults. For children, gist abstraction is of special relevance to transform new information from social and emotional contexts into stable representations. This study investigated the effect of sleep on emotional and social recognition and gist abstraction on N=34 typically developing children assessed in a sleep and a wake condition. In an emotional memory task, reward-associated stimuli were presented, while a social memory task used face-stimuli to implement social acceptance or rejection from peers. Both paradigms relied on a hidden rule to be abstracted. In general, children were able to remember emotional and social stimuli and to abstract gist information. With respect to sleep, we found a beneficial effect of sleep on the recognition of emotional stimuli but no sleep-dependent enhancement for either social recognition or emotional or social gist abstraction. Overall, our results indicate, that sleep-dependent recognition might depend on the type of memory task. Furthermore, nighttime sleep as compared to daytime wakefulness has no differential influence on gist abstraction in children as assessed in our paradigms, contrasting previous results found in adults.

## Introduction

Sleep facilitates the reorganization of memory, both quantitatively and qualitatively, through sleep-dependent memory consolidation. New information is replayed to transform memory representations stored in hippocampal regions into more stable representations in the neocortex by a reactivation of hippocampal representations in close interplay with neocortical information (Brodt et al., 2023; Diekelmann & Born, 2010; Klinzing et al., 2019; Lutz et al., 2026). The precise coordination of three oscillations during non-REM sleep supports this process: these are neocortical slow oscillations (SO), thalamocortical spindles, and hippocampal ripples (Lutz et al., 2026; Staresina, 2024).

The physiology of sleep changes across the life span, with prominent changes during childhood and adolescence. These changes comprise a decrease in the SO amplitude in older compared to younger children as well as signs of decreased REM duration and latency (Goel & Goel, 2024; Kurz et al., 2023). Changes in sleep architecture across childhood are paralleled by cortical plasticity and the evolution of memory processing (e.g. Goel & Goel, 2024; Tesler et al., 2013). For the effect of sleep on recognition memory, even though under discussion concerning memory domains and study designs (Lipinska et al., 2019; Schäfer et al., 2020), studies indicate that adults and typically developing children profit from sleep in different memory domains (e.g. language acquisition (Schimke et al., 2021; Smith et al., 2018; van Rijn et al., 2023) or episodic memory (Berres & Erdfelder, 2021; Lokhandwala & Spencer, 2021)). A recent review by Lutz et al. (2026) highlighted, that this mechanism varies with respect to different tasks or content (e.g. episodic vs. language-based memory vs. emotional memory) and might be dependent on experimental, contextual and participant-specific factors such as age or neurodivergence while some studies even show contradictory results indicating better memory performance after wakefulness. Nevertheless, for recognition of emotional and social stimuli as well as for reward-based paradigms in typically developing children studies previously reported a positive effect of sleep (Kurz et al., 2021; Prehn-Kristensen et al., 2018; Prehn-Kristensen et al., 2017; Prehn-Kristensen et al., 2013). More studies are needed that investigate sleep-associated memory processes in children, focusing on different domains and types of memory.

Aside from recognition, sleep is also assumed to facilitate qualitative changes of memory traces into schematic representations, resulting in so-called ‘gist’ information. Gist here describes the abstracted representation of reoccurring, essential information throughout various experiences (Lutz et al., 2026). While also important for adults to account for limited memory storage capacity, this is of heightened importance for children since they are exposed to a wide array of new impressions during development. The reduction and abstraction of new information, and with that the formation of gist, therefore, is crucial for their development (Mih, 2009). The definition of gist information is topic of ongoing debate. While mostly described as the product of the reduction of invariant information (like rules or categories), gist information is proposed to be an umbrella term for different, abstracted, representations, like schema or feature generalization, that are transferred to be stored in long-term memory (Dudai et al., 2015; Klinzing et al., 2019; Landmann et al., 2014; Lewis et al., 2018; Lutz et al., 2017). This abstraction of gist presumably takes place during sleep. It facilitates the adaptation to new situations based on internalized concepts extracted from prior learning situations. This process further changes during childhood, shifting from a more episodic processing to more neocortical, semantic representations (Brainerd et al., 2002; Holliday et al., 2011; Karjack et al., 2025). The establishment of higher-level cognitive representations is important for a child’s healthy development, specifically regarding their ability to adapt to social norms or adequate emotional reactions (Nicol et al., 2020). Altered social cognition, as for example in autism, can negatively impact social interaction especially in more complex and new situations (Schaller & Rauh, 2017), while differences in the emotional domain, as prominent in attention deficit hyperactivity disorder (ADHD), are associated with widespread difficulties in recognizing and processing emotions in various contexts (Soler-Gutiérrez et al., 2025). Focusing on the gist abstraction of social and emotional information in typically developing children, this study serves as a pilot study laying a foundation to investigate the role of sleep in potentially aberrant patterns of emotional and social gist abstraction in ADHD (Mirandola et al., 2012) and autism (Patry & Horn, 2019).

Currently, knowledge about the precise mechanisms of gist abstraction of emotional and social information during sleep in children is limited. There is a growing body of evidence that suggests, that the abstraction of higher-level representation of emotional and social knowledge is supported by sleep in adults (Lutz et al., 2017; Qin et al., 2022). Research on recognition and gist abstraction in children is scarce. Interestingly, in a study from Kurz et al. (2025), children showed better recognition while only adults profited from sleep when it came to the abstraction of social cues. Overall, only limited information on the abstraction specifically of emotional and social information in children is currently available. Studies investigating the influence of stimulus domain, memory type and conditions of sleep and wakefulness within the same sample are required.

Therefore, the goal of the current study was to investigate the influence of sleep or wakefulness on recognition and gist abstraction in school children using newly adapted social and emotional paradigms in a within-subjects design. For both memory tasks, social and emotional, we hypothesized better recognition memory after nighttime sleep compared to wakefulness in children. We further hypothesized greater abstraction of gist information (e.g. the ability to correctly abstract and transfer a reoccurring pattern in form of a hidden rule onto new stimuli) after sleep as compared to wakefulness.

## Methods

### Sample

Initially, 34 children participated in the study. Inclusion criteria were IQ>80, age between 8 to 12 years, sufficient knowledge of the German language, no psychiatric diagnosis and no medication (e.g. melatonin supplements) that could potentially interfere with cognition or sleep. Participants were recruited via flyers and mailing lists. One participant slept during the day while being in the WAKE-condition and therefore had to be excluded. The final sample comprised of 33 participants (15 females, mean age=10.1 ± 1.44 years). IQ, assessed with the CFT 20-R (Weiß, 2006) or the CFT 1-R (for participants under 8.5 years, Weiß & Osterland, 2013) was ≥88 for all participants. To evaluate psychological wellbeing, signs of neurodevelopmental conditions, sleep problems, and differentiating chronotype, the participants’ parents answered various screening questionnaires. The Child Behavior Checklist 6-18 R (CBCL, Döpfner et al., 2014) was used to assess general functioning and signs of various psychiatric disorders. Trait expressions of ADHD or Autism Spectrum Disorder (ASD) were assessed using the ADHD-Rating Scale (ADHD-R, DuPaul et al., 2016) and the Social Responsiveness Scale 2 (SRS, Bölte & Poustka, 2007). Further, sleep behaviors and chronotype were assessed with the Sleep Self Report (SSR, Schwerdtle et al., 2010) answered by the children and the Composite Scale of Morningness (CSM, Randler, 2014) answered by the parents. To estimate working memory capacity we used the Digit Span Task from the Wechsler Intelligence Scale-IV (WISC, Petermann & Petermann, 2011). See Table 1 for descriptive statistics. For the CSM used to assess chronotype the sample consisted of 13% evening, 36.4% morning and 60.6% intermediate type.

**Table 1.** Screening questionnaires.

| Questionnaires/Tests | Subscale<br>(if applicable) | N | Mean (M) | SE | Min | Max |
| --- | --- | --- | --- | --- | --- | --- |
| CFT 20-R /CFT 1-R | IQ max | 33 | 115.9 | 2.55 | 91 | 161 |
| CBCL | TOT | 33 | 53.0 | 1.34 | 31 | 69 |
|  | INT | 33 | 53.3 | 1.70 | 38 | 72 |
|  | EXT | 33 | 52.2 | 1.39 | 35 | 75 |
| ADHD-R |  | 33 | 9.49 | 1.28 | 0 | 33 |
| SRS | SRS-total | 32 | 47.3 | 1.58 | 26 | 69 |
|  | SRS-autism | 33 | 28.1 | 0.49 | 25 | 38 |
| SSR |  | 31 | 26.7 | 0.83 | 19 | 40 |

**Annotations:** CFT 20/1-R: IQ max: IQ value after maximal test duration; CBCL: t-values for: TOT: subscale for overall psychiatric strain, INT: subscale for internalizing problems, EXT: subscale for externalizing problems; ADHD-R: sum scores for ADHD rating scale; SRS: SRS-total: t-values for comparison with non-psychiatric age group, SRS-autism: t-values for comparison with autistic peer group; SSR: sum scores for sleep self-report.

### Study design and procedure

The study was preregistered (https://osf.io/taqs5/) and conducted as a multi-center study at the University of Tübingen as well as at the University Medical Centre Schleswig Holstein in Kiel, Germany. At both sites the study was approved by the local ethics committee of the medical faculties and conducted in line with the Declaration of Helsinki. Prior to the experimental conditions, participants and their parents gave written informed consent and underwent a screening session to ensure they meet eligibility criteria. The study was conducted as a within-subject design with each of the participants undergoing a SLEEP and a WAKE-condition (Fig. 1A). The order of the two conditions was randomized throughout participants, and they were conducted in our laboratories at least two weeks apart from each other. In the SLEEP-condition, participants arrived in the evening (approximately 3 hours before their usual bedtime), underwent testing and were instructed to go to bed shortly after. They then returned the next morning after a retention interval of approximately 12 hours to continue. During the WAKE-condition, participants came to the labs in the morning (approximately 1 hour after they usually wake up), underwent testing and then left the lab for their usual daytime activities. They then returned in the evening after a retention interval of approximately 12 hours to finish testing. Participants were instructed not to nap during the day. In both conditions, the participants underwent two tasks, an emotional and a social memory task. Task order was balanced across participants and remained the same during each condition. Each of these tasks consisted of three phases: Encoding (ENC), immediate retrieval (IMM) and delayed retrieval (DEL). Encoding and immediate retrieval were performed before the retention interval, delayed retrieval afterwards. Participants first underwent both the encoding of the social memory task and the encoding of the emotional memory task consecutively and then continued with both immediate retrieval phases after a break of approximately 20 minutes. After the 12-hour retention interval they then performed both delayed retrievals.

**Figure 1.**
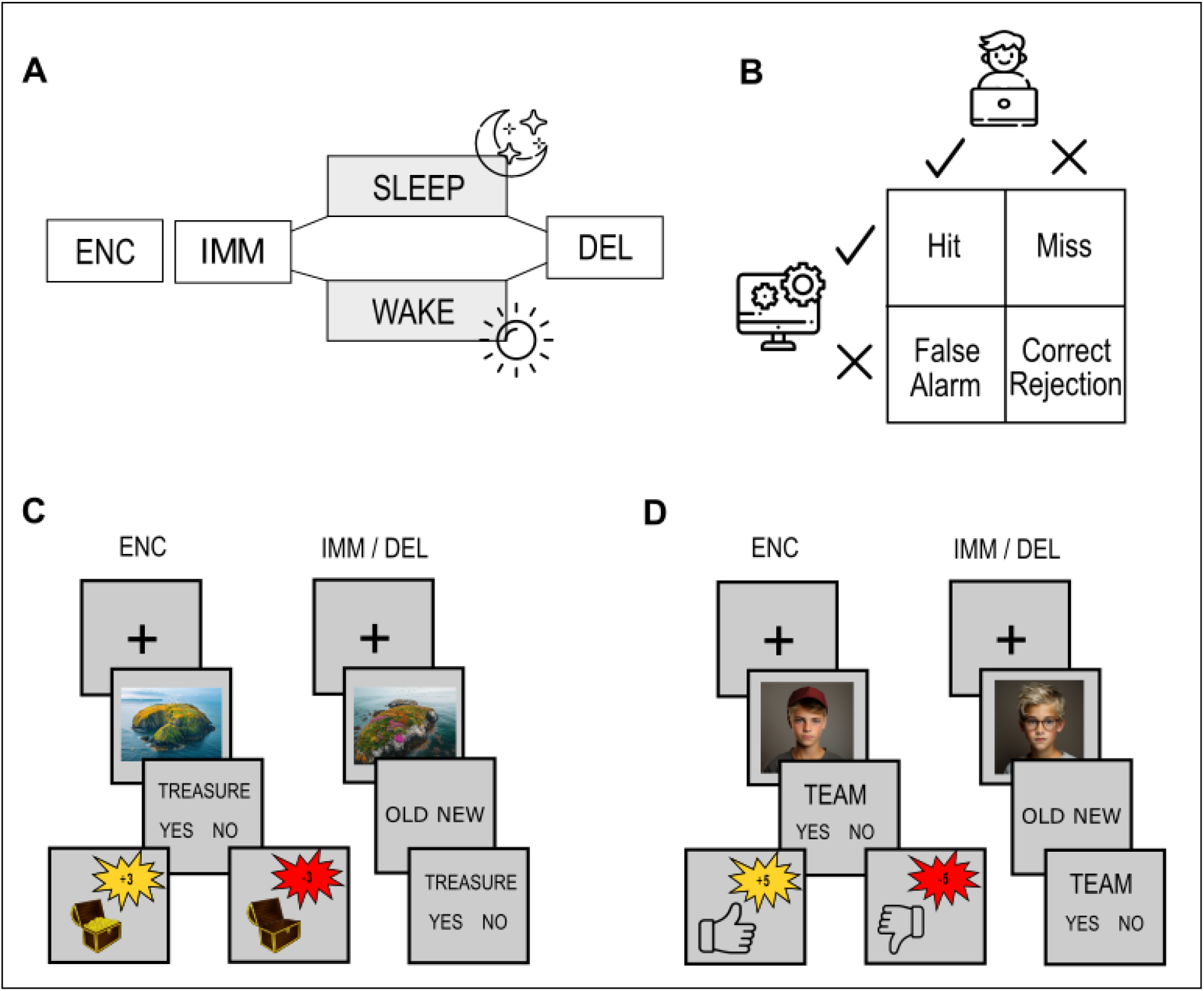
Study design and paradigms. A) study procedure. The study was conducted as a within-subject design, conditions (SLEEP and WAKE) were conducted at least two weeks apart. B) Answer options for the emotional and social memory task. C) Social paradigm. Encoding-phase on the left and retrieval phases on the right. D) Emotional paradigm. encoding-phase on the left and retrieval phases on the right. ENC: encoding, IMM: immediate retrieval, DEL: delayed retrieval.

### Stimulus material

Both tasks are based on the prisoner dilemma by Rapoport and Chammah (1965), originally proposing the situation of two separately interrogated suspects. Their behavior during the interrogation, either cooperating with each other and staying silent or defecting and betraying the other would lead to different outcomes of their sentencing: mutual cooperation would lead to moderate sentencing while mutual defection would result in a harsher punishment. But if only one of both defects, the defector would remain free of charge while the cooperator would get the harshest punishment. We adapted this paradigm to be age-appropriate and of emotional and social context for this study (Fig. 1 C&D). To provide age-appropriate stimuli that varied based on pre-defined characteristics, we used the AI MidJourney ([Midjourney](https://www.midjourney.com/home)) to generate the stimulus sets. For this study we exclusively used MidJourney‘s ‘Imagine’ prompt. An example for the prompts used in the social and emotional memory task, respectively, can be found in the supplement (Generation of stimulus material with MidJourney). For our social memory task, we generated portraits of children of about 10 years of age, for our emotional memory task photos of islands. Pictures were generated to contain target characteristics and covariables.

For the social memory task, the target characteristics were A) hats and ponytails, B) glasses and necklaces, C) scarfs and headbands and D) hoods and headphones. Variation was created by varying other characteristics such as sex, ethnicity (African American, Caucasian, Asian), and shirt color or pattern. Stimuli subsets were balanced to contain equal parts girls and boys and 50% Caucasian and each 25% African American or Asian children. The prompt further included neutral facial expression, a neutral grey background and a quality resembling a Nikon D850 with an 85mm lens. Aspect ratio remained standard MidJourney configuration to obtain square photos.

For the emotional memory task, target characteristics were A) birds and buildings, B) caves and boats, C) beaches and piers/docks and D) driftwood and agriculture. Covariates were geographical location (e.g. Pacific, Caribbean, Atlantic, Baltic Sea), vegetation (e.g. colorful, scarce, lush), weather and time of day (sunrise, sunset, daytime). The prompt further included to generate pictures that resemble a drone shot taken with a Panasonic Lumix S pro 50mmf/1.4 wide angle lens. Pictures had to have an aspect ratio of 4:3.

### Paradigms

The study comprised of an Emotional and a Social memory task that were specifically adapted for the study. We invented two age-appropriate backstories to change the context of the original paradigm to either an emotional, reward based, and a social, acceptance based, context.

In both paradigms, every phase consisted of 100 trials showing a singular picture for two seconds. Beforehand, a variable inter-stimulus interval (ISI; min. duration: 1000ms, max. duration: 1500ms) was displayed. For each paradigm, two target characteristics (see stimulus material) formed a stimulus subset, covariables varied throughout all subsets and had no predictive value. During the first condition two subsets each were presented to the participants, the two remaining subsets were presented during the following session. How the subsets were paired during conditions as well as their allocated consequence was randomized throughout participants.

### Emotional memory task

The emotional memory task encouraged participating children to imagine themselves on an expedition ship searching for hidden treasures on secluded islands (Fig. 1C). During encoding, after each stimulus presentation, participants were asked if they wanted to depart the expedition ship to enter the island and search for a treasure. Alternatively, they could stay on the ship and sail to the next island. They were instructed to keep in mind that leaving the ship to go treasure hunting would cost them 3 gold coins since an expedition would come with a certain cost. After their decision, participants were then given feedback on whether they had or had not found or missed a treasure chest, depending on whether they had entered the island or not. Feedback was given in virtual golden coins which were distributed as following (Fig. 1B): If the participant entered an island with a treasure chest on it, the participant was awarded +6 coins which made a profit of +3 coins after the expedition cost was subtracted (Hit).

If they decided not to enter an island that did not contain a treasure, they were awarded +1 coin for their correct rejection. If the participant entered an island without a treasure chest on it, they lost the −3 coins it cost to start the expedition (False Alarm). If they decided not to enter an island that did contain a treasure, the expedition company fined them −1 coin for their loss of profit (Miss). Feedback about profit or loss was accompanied by either the sound of a cash register for winning or a buzzer sound for losing coins. Unbeknownst to the participant, whether an island contained a treasure or not was determined by the target characteristics displayed in the pictures. This way, two of the four displayed target characteristics (e.g. birds and buildings) did indicate a treasure, while the other two displayed target characteristics (e.g. caves and boats) did not. Islands with and without treasures were equally distributed. Participants were further instructed to memorize the pictures, since they would be asked to remember them later.

During each retrieval phase (immediate and delayed), participants were shown 100 stimuli of which 50 had been shown during encoding while 50 were new to them. They were asked A) whether they had seen the island before (Recognition) and B) were instructed to consider whether there had been a treasure on the island or not (for OLD) or whether the island would be likely to hide a treasure or not (for NEW, Gist Abstraction). Recognition was tested through the correct identification of encoded pictures versus newly introduced pictures during both retrievals. Gist abstraction was measured for the correct application of the hidden rule for all new stimuli irrespective of whether the participant correctly identified the picture as new when asked for during the recognition. Throughout all phases, answers were given as forced choice with answer options ‘old’/’new’ or ‘yes’/’no’. Participants were not instructed to answer as fast as possible. Both answers were given self-paced. No feedback was given either during immediate or delayed retrieval.

### Social memory task

In the social memory task, participants were instructed to imagine themselves in a summer camp where they had to choose teammates for a group activity (Fig 1D). During encoding, participants were asked if they wanted the presented child to be on their teams or not. They were told beforehand that after they had accepted or denied one of the displayed children, said child would ‘decide’ if they themselves would ‘want to be on their team’ or not. Participants were then given feedback on how the portrayed child had decided. As in the emotional memory task, this led to four possible scenarios (Fig. 1B): if both the participant and the stimulus wanted to form a team the participant was awarded +5 points (Hit). If they both declined being on a team with each other, the participants were awarded +2 points for their correct rejection. If the participant did not choose the displayed child despite the child wanting to make the team −2 points were withdrawn (Miss). If the participant chose to invite the displayed child to be on his team but was rejected −5 points were withdrawn (False Alarm). Whether the stimulus would accept or reject the offer was determined by target characteristics displayed in the picture. Again, two target characteristics indicated acceptance and two rejection, consequences were distributed equally and participants were again instructed to memorize the pictures.

During both immediate and delayed retrieval, participants were asked A) whether they had seen the child before (Recognition) and B) whether said child previously wanted to be on their team (for OLD) or would be likely to accept their offer (for NEW, Gist Abstraction). Answers were given self-paced and participants did not receive feedback.

### Control measures

As control measures, before each phase, participants performed a psychomotor vigilance test (PVT, Basner et al., 2011) to assess vigilance. Further, they were asked about their mood (self-developed 5-point scale from very bad to very good), their motivation (self-developed 5-point scale from no motivation to high motivation) and their current level of sleepiness using the Stanford Sleepiness Scale (SSS, Hoddes et al., 1972). Significant differences, regarding vigilance, mood, and motivation might result in limited interpretability of our main experimental data.

### Statistical Analysis

Statistical analysis was performed in R version 4.4.2 (R Core Team, 2024). Recognition memory and gist abstraction performance were calculated using d-Prime defined as d’ = z(Hits)-z(False Alarms). D-Prime for Gist Abstraction is defined as difference of the correctly associated consequence for all NEW items, even those that participants falsely remembered as OLD (“new” + “old” | NEW) minus the False Alarms. While originally preregistered as only those NEW items that had been correctly identified as such due to participants having different amounts of “new” decisions, we decided to calculate d-prime values for gist abstraction based on consequence decisions for all new items, irrespective of the participants’ old/new decision. Control analysis revealed, that this did not influence the outcome of our study.

We originally pre-registered to compare the performance in the sleep and wake conditions using paired t-tests by calculating the difference between the immediate and delayed phase. We opted against the difference score and included the fixed effect Retrieval (immediate vs. delayed) for a better interpretability of effects. To assess recognition and gist abstraction in both tasks, we used two linear-mixed effects models using the R package lme4 (Bates et al., 2015). We used the library lmerTest (Kuznetsova et al., 2017) to compute F-statistics and p-values using Satterthwaite’s Method and Type III sums of squares. Post-hoc analysis was conducted using the emmeans package (Lenth, 2023). The linear mixed models to calculate gist as well as recognition included the fixed effects Condition (sleep vs. wake), Retrieval (immediate vs. delayed) and their interaction terms, as well as participants as random intercept. The model therefore was defined as:

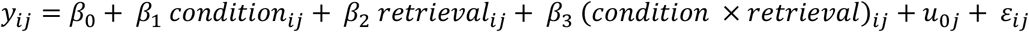

To control for multiple comparisons, we corrected p-values for each model using the Bonferroni method.

For the analysis of control measures Wilcoxon Rank tests were performed for ordinal data obtained from questionnaires regarding mood, motivation and sleepiness and t-tests for the PVT to control for differences between conditions and phases. Further, the difficulty of the emotional and the social memory task was compared using a linear mixed model with the factor Task (emotional vs. social) alongside the factors Condition and Retrieval. We did not proceed with originally pre-registered correlational analysis of screening questionnaires on ADHD and autism trait expressions. The data of these questionnaires, since collected in children without psychiatric diagnosis, lacked necessary heterogeneity.

Unless otherwise noted, data will be reported as mean ± standard error of the mean (SE).

## Results

### Recognition

For recognition in both tasks, statistical analysis of d-Prime values across all phases revealed recognition above chance-level (all p<.001; for further information see supplement Table S1) indicating that the children were able to remember the previously encoded items in both tasks.

For the emotional gist task (Figure 2A), there was a significant interaction for retrieval x condition (F(1,99)=8.68 p=.004). Follow-up tests revealed a decrease of recognition from immediate to delayed in the wake condition (M=-0.34±0.002) while it remained stable in the sleep condition (M=0.07 ± 0.01) The was a significant main effect for retrieval (F(1,99) 18.83, p<.001, IMM: M=0.587±0.056; DEL: M=0.382±0.048), but not for condition (F(1,99)=0.59, p=.445, sleep: M=0.466±0.053; wake: M=0.502±0.051).

**Figure 2.**
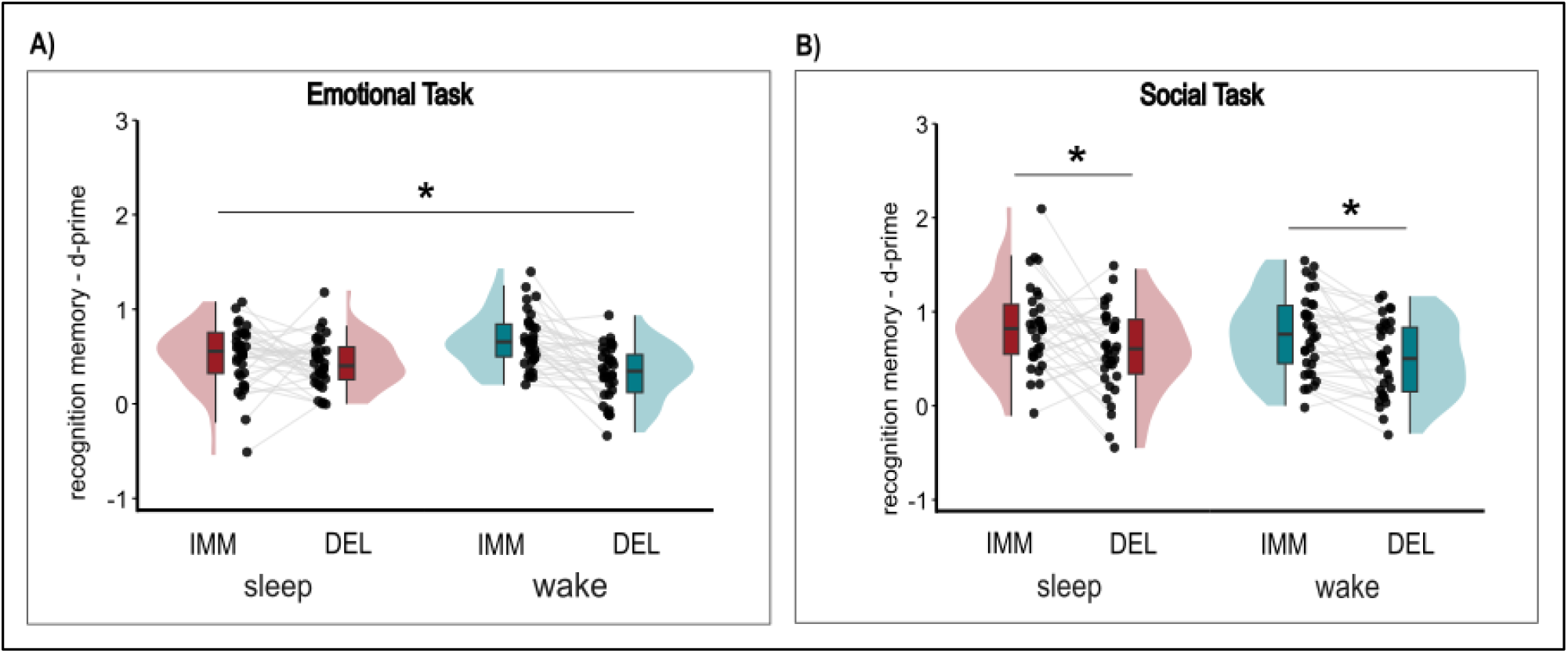
Results for Recognition. Participants showed greater forgetting in the wake compared to sleep condition in the emotional task and an overall decrease in recognition from immediate to delayed retrieval in the social task. Recognition performance (d-Prime) for an A) emotional memory task and B) social memory task. IMM: immediate retrieval, DEL: delayed retrieval.

For the social memory task (Figure 2B), there was no significant interaction for condition x retrieval (F(1,96.8)=0.0014, p=.969). However, there was a significant main effect retrieval (F(1,96)=19.84, p < .001) with worse recognition performance during delayed (M=0.525±0.005) compared to immediate retrieval (M=0.802±0.055). There was no condition main effect (F(1, 97.93)=1.32, p =.253, sleep-wake; M=0.07±0.006).

For both tasks, participants showed significantly better performance in the second session of the within-subject design independently of whether they had been through the sleep or the wake condition for the first session. The addition of the factor Session (first time performing the tasks vs. second time performing the tasks) into the linear mixed model on exploratory level showed a highly significant ‘Session x Retrieval’ interaction effect indicating better recognition in the immediate retrieval of the second compared to the first time performing the tasks during the within-subject design (Session x Retrieval interaction for Emotional memory task: F(1,99)=12.0230, p=.001 and social memory task: F(1,96.765)=6.54, p=.012). The significant condition x retrieval-interaction in the emotional memory task was not affected by this session effect.

To determine whether both tasks differed in terms of recognition difficulty, the recognition data for the two tasks were compared with each other. We operationalized difficulty as the participants’ performance indicated by d-Prime. These analyses showed significantly higher d’ values in the social memory task (M=0.662±0.039) compared to the emotional memory task (M=0.484±0.028; main effect task: F(1,228.87)=18.43, p<.0001), indicating an overall higher difficulty of the emotional memory task when it comes to correct remembrance. For differences regarding task difficulty during immediate and delayed retrieval as well as the interaction of task and retrieval see the supplement (Table S2).

### Gist abstraction

Children were generally able to extract gist information in both tasks as indicated by their above chance-level performance during encoding as well as retrieval (all p < .001, see Supplement Table S3 for further information).

For gist abstraction in the emotional memory task (Figure 3A), results revealed neither a significant interaction of condition and retrieval (F(1,99)=0.09, p=.769), nor significant main effects for condition (F(1,99)=0.35, p=.556, SLEEP-WAKE, M=0.083±0.005) or retrieval (F(1,99)=0.01, p=.933; IMM-DEL, M=-0.012±0.007).

**Figure 3.**
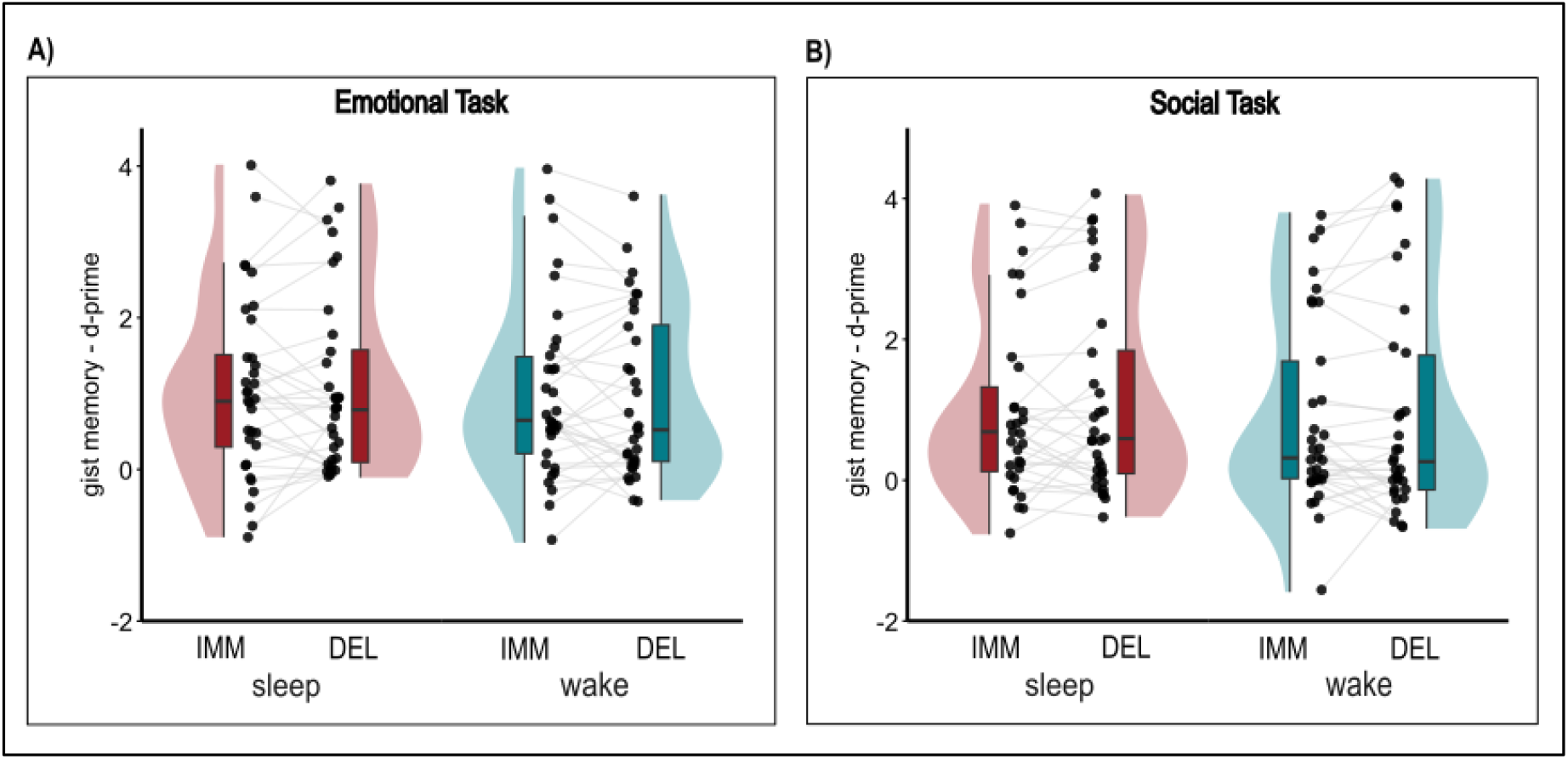
Results for Gist abstraction. Participants showed no sign of differentiating gist abstraction performance (d-Prime) for an A) emotional memory task and B) social memory task with respect to sleep or wakefulness. IMM: immediate retrieval, DEL: delayed retrieval.

Similar results can be reported for the social memory task (Figure 3B): Results showed neither a significant interaction of condition and retrieval (F(1,95.6)=0.05, p=.822), nor significant main effects for condition (F(1,96.455)=0.78, p=.379; SLEEP-WAKE; M=0.151±0.01) or retrieval (F(1,95.6)=0.23, p=.630; IMM-DEL; M=-0.116±0.0021).

When session was added as a factor to the model then a significant main effect (Emotional memory task: F(1,99)=23.28, p < .001; social memory task: F(1,96.441)=20.47, p < .001) occurred showing that gist abstraction was better during the second session compared to the first session in both paradigms.

As for recognition, we further analyzed task differences regarding the difficulty of gist abstraction. We therefore analyzed the performance during immediate and delayed retrieval (see Supplement Table S2) and furthermore the correct identification of pictures with positive and negative consequence during encoding (see Supplement Table S4). Results this time did not reveal any significant differences for task, retrieval or their interaction (p ≥ .301). Further, exploratory analysis revealed a significant effect of task order during encoding which is explained further in the supplement (see Supplement Table S5).

### Control measures

The analysis of the control measures motivation, sleepiness and mood revealed, that the participants were less motivated during delayed retrieval in the wake condition (in the evening, M=3.85±0.16) than during the sleep condition (in the morning, M=4.24±0.12; Wilcoxon’s z=2.42, p=.016) but not during encoding or immediate retrieval. Adding motivation (during immediate and delayed retrieval) as a covariate to the models for recognition and gist abstraction did not change any of the reported results. Mood and sleepiness did not differ significantly between any of the phases or between conditions (p ≥ .102). Psychomotor vigilance assessed with the PVT revealed no differences regarding phase or condition as well (p=.866). Due to the self-paced nature of the participants’ answers in the paradigm, we did not analyze reaction times.

## Discussion

In the present study, we investigated whether sleep has a positive effect on recognition memory and gist abstraction of emotional and social information in children. Only for the emotional task, we did see a positive effect of sleep on the recognition performance resulting in less forgetting in comparison to the wake condition. While both tasks were able to assess gist abstraction in children in general, we did not find any influence of sleep on gist abstraction.

Concerning the recognition effect, that we found for the emotional memory task, these findings integrate well into findings from previous studies showing that sleep in children tends to prevent greater forgetting compared to wakefulness (Kurz et al., 2025) and specifically effects the recognition of emotional, reward based information (Kurz et al., 2023; Prehn-Kristensen et al., 2018). Further, greater loss of memory during the wake condition compared to the sleep condition goes in line with a study from Fenn and Hambrick (2013), who indicate that sleep might protect new memory from loss rather than to add its enhancement. Contrasting these findings, we could not show the same effect for our social paradigm, differing from earlier, predominantly physiological results from Prehn-Kristensen et al. (2017), where children profited from sleep when performing a social task instructing them to memorize faces of differentiating emotional valence. Possibly these differences result from the stimulus material: while the current study focused on a group based acceptance/rejection-cue using emotionally neutral faces, Prehn-Kristensen et al. (2017) used faces displaying different emotions that had to be rated in their perceived emotional quality. This resulted in a sleep effect mainly driven by the faces perceived as angry with general differences regarding different facial expressions, possibly indicating that facial expression might play a crucial role in the remembrance of faces. Our results, therefore, underline the heterogenous set of findings concerning a sleep effect on recognition as described by Lipinska et al. (2019) and highlight the need to further research what mechanisms lead to improvement of recognition fostered by sleep in children.

For gist abstraction other than hypothesized, neither during the social nor during the emotional memory task, our participants showed a significant improvement after sleep. While d-Prime values significantly above chance level indicate that children indeed abstracted some underlying gist information during the experiment, we could not show any difference resulting from the condition, the overall delay between immediate and delayed retrieval or their respective interaction. Furthermore, our participants already showed d-Prime values significantly above chance level during encoding, indicating they were already able to attribute the correct consequence to the picture while still learning and therefore already abstracted gist. This shows the general ability to rapidly abstract gist, but it also highlights that sleep or wakefulness right after learning might not play a differential role here – this is supported by findings from Lutz et al. (2025), who concluded rapid gist abstraction seems to be independent of sleep or wakefulness afterwards and sleep only improves performance after substantial durations. Our results therefore go in line with previous findings from Kurz et al. (2025) who did not find greater feature generalization after sleep in declarative memory tasks but contrast results indicating children’s better abstraction in motor memory tasks (e.g. Wilhelm et al., 2012; Wilhelm et al., 2013) and more so findings from studies with adult participants indicating gist abstraction during sleep for various domains of memory (e.g. Lutz et al., 2017, Lutz et al., 2026).

To explore whether task difficulty might have affected recognition and gist abstraction performance, we examined the participants’ performance measured with d-Prime. Results for recognition memory indicate significantly lower d-Prime values for the emotional memory task compared to the social memory task. This might propose an explanation as to why there is a sleep effect for recognition in the emotional memory task and not in the social memory task. If children struggled more to remember pictures from the emotional memory task overall this might explain how during sleep those less stable memories might have been protected through memory consolidation while children tended to forget more during wakefulness. It has to be highlighted here that neither task difficulty was explicitly manipulated nor was task specific difficulty systematically assessed in any other way. Still, different studies highlight task complexity or encoding strength as a possible influence on overnight consolidation. One can argue less efficiently encoded or more complex stimuli in different memory domains (e.g. Blischke & Malangré, 2017; Petzka et al., 2021; Schapiro et al., 2018) might propose an interesting starting point to further investigate the kind of information that profit from memory consolidation during sleep. Lastly, our results show a dominant session effect. For both recognition and gist abstraction in each task, we see significantly better performance in the second session. Given that the two experimental sessions were at least two weeks apart from each other, improved performance might result from the bypassing time. Several studies noted that an only 12-hr long retention interval might not be enough time for memory consolidation and gist abstraction to support memory performance and the reduction and transformation of memory might take longer (e.g. Dudai et al., 2015; Lutz et al., 2025). In our specific case it is important to note that during the second session we presented entirely new stimuli, but the procedure and task structure remained the same. So, while participants were confronted with new stimuli the prior knowledge of the task requirements and knowledge about an underlying rule as gist information might have improved their performance.

## Limitations and future perspective

The interpretability of our results has some limitations: First, originating from our within-subject design, our data was strongly affected by a significant session effect. Children performed significantly better in the second session independently of the condition. The comparability of post-encoding sleep and wakefulness therefore might be constrained by the significance of the session-factor. We assured our significant recognition effect in the emotional memory task remained as such, when incorporating session as a factor into our analysis. Still future studies might either profit from a between-subject design or longer between-session intervals.

Further limitations pertain to the emotional and social context of our paradigm: For the emotional paradigm, while reward-associated learning is widely seen as part of emotional memory processes (e.g. Diekelmann et al., 2009), our stimuli themselves were of no emotional value. In the social paradigm, the shown portraits of children with their acceptance or rejection of the participants might be feasible to create social context but it remains unclear if fictional children in a fictional social situation are of enough salience to be preferably consolidated overnight (Alger et al., 2019). It would be interesting to investigate whether the proposed monetary reward of the emotional memory task’s expedition backstory provides different incentives compared to the points rating social acceptance in the social task’s summer camp backstory. Even though participants were not actually rewarded with money from the paradigm, proposed monetary reward might have improved saliency of the emotional stimuli and might be specifically fostered by post-encoding sleep compared to wakefulness (Lutz et al., 2026; Prehn-Kristensen et al., 2020; Sterpenich et al., 2021) while the social context of the summer camp might fall short behind and be therefore of lower saliency for memory consolidation during sleep. To at least partly control for this effect, participants were instructed to memorize stimuli for a later recall. Still we do not know how this influences the memorization and consolidation of the emotional and social value. Taken together with possible differences in difficulty, possibly fostering an altogether higher consolidation of the more difficult and more salient emotional stimuli it remains unclear if the social stimuli would profit from sleep, if they weren’t overshadowed by their emotional counterparts. Ultimately, we cannot directly compare the different paradigms to clearly argue for effects of stimulus content. Future paradigms might try to parallelize emotional and social conditions or to the contrary opt for very distinct task designs to avoid unclear relation between stimulus material and task design.

Possible future study designs could focus on incorporating solely social stimuli and no parallel emotional task or enhance the salience of perceivably neutral stimuli like plain faces by adding a more emotional context to them, for example by presenting them with emotional attributions (Medford et al., 2005) or presenting them alongside non-social cues (Grunewald & Schweizer, 2026). Another hypothesis might be, that faces are of such high salience for our everyday life that they are consolidated overnight but probably also underly rapid consolidation processes in the first 4 hours after encoding (Wiese et al., 2024). This highlights the need to further research different consolidation processes of different memory domains, and the role sleep might play in it.

Another aspect that is left unaddressed in this study is the role of age and development. With participants in the ages of 8 to 12 years, the study focuses on a narrow range developmental activity. It might possibly be that the formation of schemata has been conducted earlier in life, leaving less flexibility to abstract new gist information (e.g. Hartley et al., 2021). On the other hand, it remains unclear whether aging would possibly have a different effect. Adults might be more sensitive to social cues because of their relevance in everyday life and show differing patterns of memory consolidation than children (Kurz et al., 2025; Prehn-Kristensen et al., 2018). Other model populations to investigate memory consolidation and especially gist abstraction during sleep might be neurodevelopmental conditions, such as children with ADHD or autism (Kurz et al., 2019; Prehn-Kristensen et al., 2013). Previous studies have shown that in these populations, recognition as well as the abstraction of information after sleep differ from typically developing children. Future studies, therefore, should not only focus on specific processes in memory consolidation during sleep but should also take development and ageing into account.

## Conclusion

The study investigated the influence of sleep on the consolidation and gist abstraction of emotional and social stimuli. While sleep stabilized recognition rates for emotional stimuli compared to greater memory loss in wakefulness, there were no signs of a beneficial influence of sleep on gist abstraction in an emotional or social context, as well as no beneficial effect for the recognition of social stimuli. Further exploratory results suggest gist abstraction already started at encoding. Our study therefore highlights that a beneficial effect of sleep on recognition memory depends on memory contents, in our case favoring emotional, reward-based stimulus material. Furthermore, our results indicate that gist abstraction as a rapid process already begins during encoding and paradigms have to carefully adjust for difficulty to leave room for improvement during the retention interval. More so, we need to discuss study procedures when initiating gist abstraction to prevent carry-over effects in within-subject designs. The study therefore underlines that future research on the one hand should investigate the recognition of different kinds of memory content, and on the other hand has to carefully develop ways to facilitate gist abstraction in a way that allows the investigation of consolidation processes afterwards. We have to highlight here that the limitations of this study do not allow for an interpretation of no beneficial effect of sleep on gist abstraction but highlight the need to further investigate the formation, mechanisms and deviations of emotional and social gist abstraction.

## Data Availability

All data produced in the present study are available upon reasonable request to the authors

## Acknowledgements

The study was supported by a grant from the Deutsche Forschungsgemeinschaft (468645090).

