## supplement Table S1 for "The influence of sleep on emotional and social recognition memory and gist abstraction in children"

**Supplement to Meyer-Jajkov et al. (2026)**

### Generation of stimulus material with MidJourney

Stimuli for both tasks were generated using the AI MidJourney. For the emotional task the stimuli were generated with prompts like the following:

> *"a photo of a secluded Island in the {pacific, Caribbean, North Sea} with {dense, colorful, scarce} vegetation with a {sailboat, cargo ship, motorboat, paddleboat} in front of it, drone shot from a low angle, Panasonic lumix s pro 50mmf/1.4, --ar 4:3".*

For the social task a prompt was constructed as following:

> *"a photo of a young {afroamerican, asian, caucasian} {boy, girl} with glasses in front of a light grey backdrop, Close up shot::5, neutral facial expression ::4, natural light, neutral white background, Nikon D850, 85mm lens".*

Covariates such as vegetation or time of day for the emotional task and ethnicity or shirt color for the social task were varied across prompts to diversify the outcome.

### Tables

**Table S1.** Recognition d-Prime values tested against chance level (t-test against zero, two-tailed), separately for the emotional and social task for both retrievals.

| | | immediate retrieval | | | | | delayed retrieval | | | | |
|---|---|---|---|---|---|---|---|---|---|---|---|
| | | *t* | *df* | *p* | *M* | *SE* | *t* | *df* | *p* | *M* | *SE* |
| emotional | sleep | 8.44 | 32 | **<.001** | 0.499 | 0.059 | 9.31 | 32 | **<.001** | 0.433 | 0.046 |
| | wake | 12.92 | 32 | **<.001** | 0.674 | 0.052 | 6.60 | 32 | **<.001** | 0.33 | 0.05 |
| social | sleep | 10.04 | 30 | **<.001** | 0.841 | 0.084 | 7.25 | 31 | **<.001** | 0.559 | 0.077 |
| | wake | 10.47 | 32 | **<.001** | 0.765 | 0.073 | 7.11 | 32 | **<.001** | 0.491 | 0.069 |

**Table S2** Comparison of task difficulty (indicated by d-Prime) between emotional and social task during retrievals, separately for recognition and gist abstraction.

| | recognition | | | | gist abstraction | | | |
|---|---|---|---|---|---|---|---|---|
| | *F* | *NumDF* | *DenDF* | *p* | *F* | *NumDF* | *DenDF* | *p* |
| Condition (sleep/wake) | 0.159 | 1 | 228.87 | .69 | 0.979 | 1 | 228.05 | .324 |
| Retrieval (imm/del) | **33.412** | **1** | **228.36** | **<.001** | 0.118 | 1 | 227.88 | .731 |
| Task (social/emotional) | **18.425** | **1** | **228.87** | **<.001** | 0.175 | 1 | 228.05 | .676 |
| condition x retrieval | 2.64 | 1 | 228.36 | .106 | 0.142 | 1 | 227.88 | .706 |
| condition x task | 1.602 | 1 | 228.87 | .207 | 0.068 | 1 | 228.05 | .794 |
| retrieval x task | 0.763 | 1 | 228.36 | .383 | 0.201 | 1 | 227.88 | .654 |
| condition x retrieval x task | 2.915 | 1 | 228.36 | .089 | 0.000 | 1 | 227.88 | .989 |

Note. imm = immediate retrieval, del = delayed retrieval; A main effect of task revealed better recognition performance in the social compared to the emotional task. Across tasks, recognition was better during the immediate compared to the delayed retrieval. No effect of task was seen regarding gist abstraction.

**Table S3.** Gist d-Prime values tested against chance level (t-test against zero, two-tailed), separately for the emotional and social task at encoding and both retrievals.

| | | encoding | | | | | immediate retrieval | | | | | delayed retrieval | | | | |
|---|---|---|---|---|---|---|---|---|---|---|---|---|---|---|---|---|
| | | *t* | *df* | *p* | *M* | *SE* | *t* | *df* | *p* | *M* | *SE* | *t* | *df* | *p* | *M* | *SE* |
| emotional | sleep | 6.48 | 32 | **<.001** | 1.146 | 0.177 | 5.1 | 32 | **<.001** | 1.056 | 0.207 | 5.35 | 32 | **<.001** | 1.085 | 0.203 |
| | wake | 5.65 | 32 | **<.001** | 0.981 | 0.162 | 4.93 | 32 | **<.001** | 1.014 | 0.206 | 5.05 | 32 | **<.001** | 0.961 | 0.19 |
| social | sleep | 3.94 | 31 | **<.001** | 1.002 | 0.225 | 4.25 | 30 | **<.001** | 0.966 | 0.227 | 4.61 | 31 | **<.001** | 1.153 | 0.25 |
| | wake | 4.09 | 32 | **<.001** | 0.837 | 0.205 | 3.74 | 32 | **<.001** | 0.887 | 0.237 | 3.45 | 32 | **<.001** | 0.932 | 0.27 |

**Table S4.** Comparison of task difficulty (indicated by d-Prime) between emotional and social task during gist abstraction at encoding.

| | encoding | | | |
|---|---|---|---|---|
| | *F* | *NumDF* | *DenDF* | *p* |
| condition | 1.081 | 1 | 98.12 | .301 |
| task | 0.404 | 1 | 98.12 | .526 |
| condition x task | 0.939 | 1 | 98.12 | .843 |

Note. There was no difference between the social and emotional task regarding the rule abstraction performance during encoding.

**Table S5.** Linear Mixed Model comparing rule abstraction performance in the first session during encoding of the social and emotional task, depending on which task was performed first within the first session.

| | encoding | | | |
|---|---|---|---|---|
| | *F* | *NumDF* | *DenDF* | *p* |
| Condition (sleep / wake) | 0.168 | 1 | 33.065 | .684 |
| Task (social / emotional) | 1.696 | 1 | 32.981 | .201 |
| task order (1 / 2) | 1.5 | 1 | 38.934 | .228 |
| condition x task | 0.327 | 1 | 32.981 | .571 |
| condition x task order | 0.355 | 1 | 38.934 | .555 |
| **task x task order** | **6.393** | **1** | **35.665** | **.016** |
| condition x task x task order | 3.03 | 1 | 35.665 | .09 |

Note. Concerning rule abstraction during encoding of the emotional task compared to the social task in the first session, the significant interaction indicates that participants performed better if the emotional task was performed after the social task, resulting in higher d-Prime values. Direct pairwise comparisons did not indicate better performance in any task when either the social or emotional task were performed first.
